# Assessments of heavy lift UAV quadcopter drone to support COVID 19 vaccine cold chain delivery for indigenous people in remote areas in South East Asia

**DOI:** 10.1101/2021.01.09.21249494

**Authors:** Andrio Adwibowo

## Abstract

Vaccine delivery is one important aspect need to be strengthened within health systems. One of the main challenges in COVID 19 vaccine delivery is how to cover indigenous population in remote and isolated forests in South East Asia. Another issue in COVID 19 cold chain delivery is requirement for a carrier that can maintain the suitable storage temperature. Related to this condition, COVID 19 vaccine should be delivered using heavy vaccine cooler box and this demand delivery system equipped with heavy lift capacity. In here, this study proposes and assess the potential used of heavy lift UAV quadcopter to expand the COVID 19 vaccine delivery to indigenous people living in village that impeded by rugged terrain. The landscape and terrain analysis show that access to the villages was dominated by 15%-45% slopes and the available access is only 1.5 m width trail. To transport 500 vials with 10 kg carrier along 2 km trail, it requires 2 persons to walk for 1 hour. By using drone, a straight line route with a length of 1.5 km can be developed. There were at least 3 drone types were available commercially to lift 10 kg load and several drones with payload capacity below 10 kg. For carrying 100 vials to village using drones, it is estimated the required delivery time was 1.23-1.38 minutes. Around 1.57-1.66 minute delivery times were required to transport 250 vials. For carrying the maximum and full loads of 500 vials or equals to 10 kg load, a drone requires in average of 3.13 minute delivery times. This required drone delivery time is significantly below the required time by walking that almost 1 hour. Drones were limited by flight operational times. Whereas all required delivery times for each drone assessed in this study were still below the drone operational time. The lowest drone operational time was 16 minutes and this is still higher than the time required for a drone to deliver the vaccine. Considering the effectiveness and anticipating vaccine vaccination, UAV quadcopter drone is a feasible option to support COVID 19 vaccine delivery to reach indigenous people in isolated areas.

## 1 . Introduction

The COVID 19 pandemic after 1 year has entered a new phase. In the midst of new emerging COVID 19 variants and large numbers of countries are still struggle with the steady increasing COVID 19 cases, now it is the time to focus on the vaccination efforts. Important issues regarding COVID 19 vaccination is related to the distribution mechanism known as cold chain delivery. COVID 19 vaccine requires specific endless low temperature storages. Any COVID 19 vaccine will need to be handled within specified temperature ranges that are likely to follow those for the influenza vaccine, which must be kept between 2°C and 8°C, while both in transport and storage facilities (Wang et al. 2020).

The second issue is related to the distribution mechanism. Since COVID 19 has infected all people whether in mountain or in coast from Antarctica to Sahara desserts, then the cold chain delivery should cover all those areas. A platform that can do that arduous logistic task is undoubtedly known as unmanned aerial vehicle (UAV) or commonly known as drone. Since the start of the pandemic, drones have played a critical role in maintaining public safety. Their roles include conveying stay-at-home messages in Dubai, washing streets in Ahmedabad, India and providing contactless delivery in Wuhan, China.

As demands for rapid COVID 19 vaccine distribution, questions remain over how the vaccine can be distributed and reached rural areas to deliver life-saving treatment or preventative inoculation. In April in Ghana, the drones have pioneered and supported COVID 19 logistics. At that time, drones have been used to deliver packages containing blood samples to rural hospitals located up to 85 kilometers away.

South East Asia should be a region that starts to use drone assistance especially for supporting COVID 19 cold chain delivery. First, countries in SE Asia were formed due to the volcanic activity and this causes the rugged terrain presences in SE landscapes (Sidle et al. 2006). The rugged terrain in the form of high mountain and deep valley with poor land transportation networks will hinder or even make it impossible the COVID 19 vaccine delivery.

The rugged terrain landscape has positive consequence in the form of fertile lands. The presence of this land that very suitable has led to the existences of indigenous people living and inhabiting remote forests. Indonesia is one of country in SE Asia that rich with cultural diversity including indigenous people with almost 70 million indigenous peoples living from city, suburban to deep in the forest. Those Indonesia’s indigenous peoples also face further challenges in addition to COVID 19. Vulnerability of those indigenous people has required them to be also prioritized in vaccination. Whereas, regarding most indigenous people reside deep in the remote forest, an approach to support the COVID 19 vaccine cold delivery is required immediately. In here, this paper is aimed to assess the feasibilities of various drone specification for supporting COVID 19 vaccine cold deliveries.

## 2 . Methodology

Methods used in this study include the assessment of landscape and analysis on drone specifications. The landscape data were collected from remote sensing analysis and drone specifications from drone manufacturer database.

### Study area

The study area is Baduy village, Banten province, Indonesia (Figure 1). Baduy is indigenous people group living in West Java Indonesia. They are living in several traditional villages inside the forest reserves. A village that assessed in this study was a village that located at outer boundary of Baduy. The population of Baduy in this village was 10486 people.

**Figure 1.**
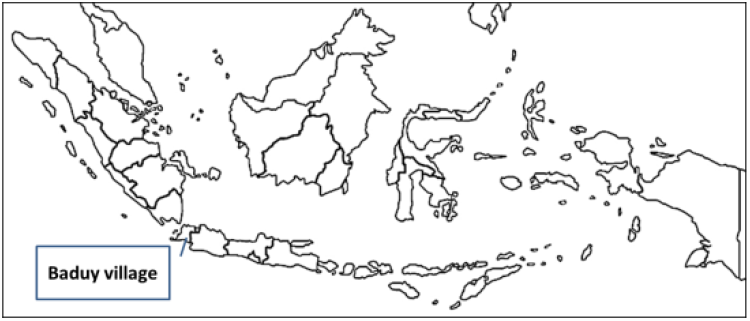
Location of Baduy village.

### Landscape assessment

Landscape assessment is required since delivery using land transport is assessed. The assessment includes the available and dimension of trails (length and width), obstruction include the presence of landslide and flash flood, and slopes denoted in percentage. Steep trail indicated by high slope % will affect and slow the delivery.

### Land transport assessment

Land transport assessed in this study includes the time required to travel to the village from the main road. The assessments include the time required to travel either by walking or using vehicles.

### Vaccine carrier specification

Vaccine carrier is the load for drone. The cold carrier box was able to maintain vaccine at low temperature for 3 days. The dimension of carrier is 43 cm x 30 cm x 30 cm. The total weight of the carrier was 10 kg and held 500 x 0.5 ml vials.

### Drone specification

The studied drones were the drone that has ability to carry heavy load >10 kg following the vaccine carrier weight. Several available commercial drones were sampled. The basic assessed drone parameters include speed (km/h) and maximum operation times (minute).

## 3. Results

### Landscape assessment

The Baduy village was located 2 km from the main road that can still accessed by the vehicles. The only trail available to access the village is a 1.5 m width track. The Baduy village was located inside the valley (Figure 2) and surrounded by high hills with slope ranged from 15-45% and it is quite steep in some locations. The village height was 220 above the sea levels (asl) and the highest hill was 342 m asl. The trails passed rivers and hills. During rainy season, the rivers have risks to cause flash flood and landslide risk due to the presence of the hills. In rainy season, the tracks can be very slippery.

**Figure 2.**
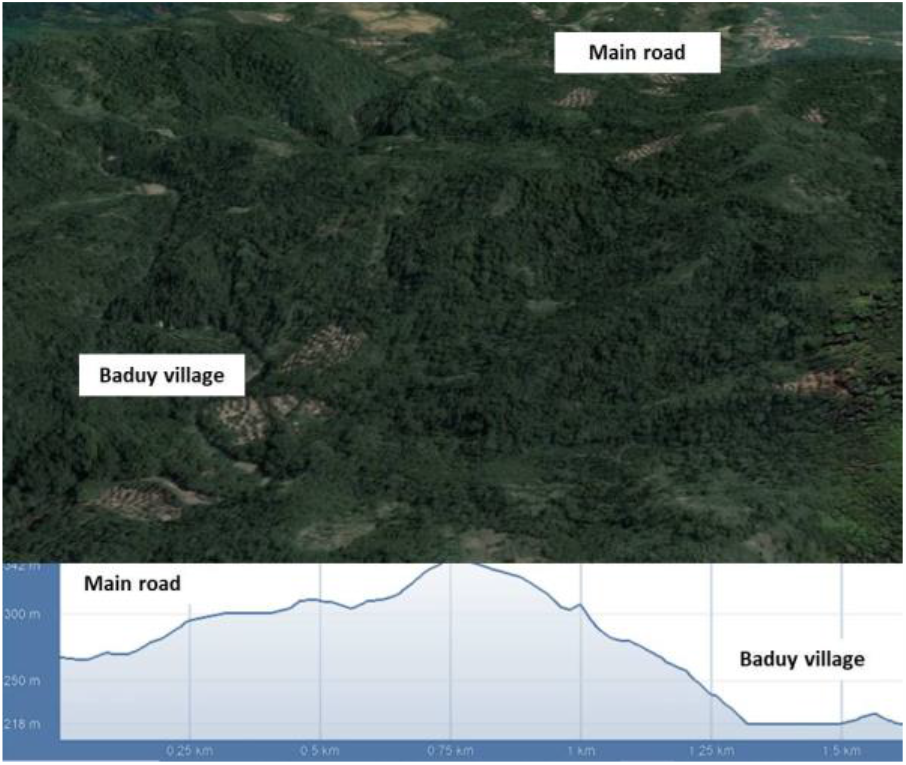
Landscape, terrain and slope of Baduy village.

### Land transport assessment

The land transport assessment shows that travelling carrying vaccine cooler box by walking from main road to the village requires 60 minutes (Figure 3). Whereas, travelling using vehicle requires 30 minutes. Whereas the transport was not in smooth and flat routes since the village was located in the valley. To reach the village, the walking transporter should pass and climb hilly areas with 15-45% slope and rivers. Then to transport 500 vials require 60 minute by walking.

**Figure 3.**
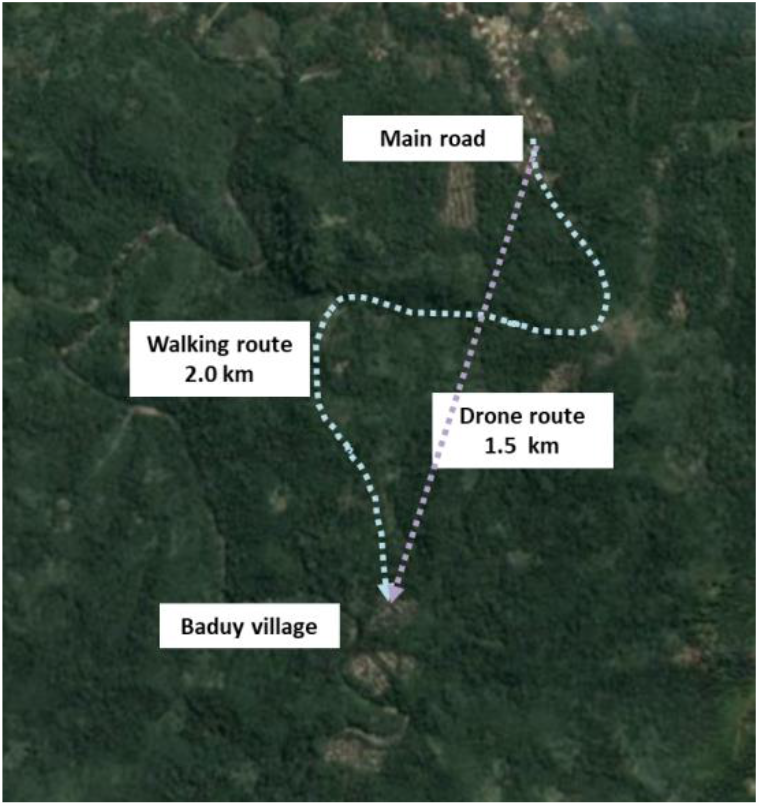
Walking and drone routes to access Baduy village.

### Drone transport assessment

Different to the land transport, the drone transport was possible conducted in straight line routes. The length of the drone transport route was 1.5 km (Figure 3). The drone transport does not require to go through up and down hilly routes.

There were 8 heavy lift drones available for commercial used (Figure 4). The assessed drones were UAV quadcopters. The load capacity was ranged from 3 to 20 kg and the range for flight time was 16-50 minutes.

**Figure 4.**
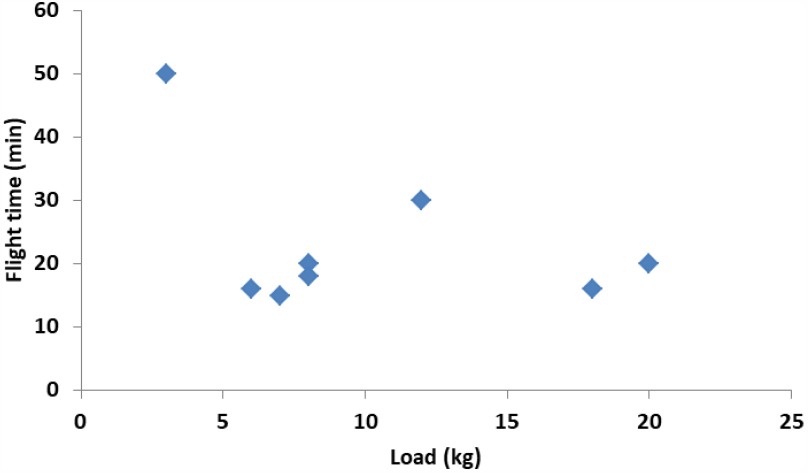
Drone payload and flight time.

Various drone types have specific load and flight operational time. Figure 4 shows the negative correlations between loads and flight times. It indicates that flight time was limited by the load. From the drone assessed, there were several drones that have payload below 10 kg and others were above 10 kg. The drone types that able to carry load larger than 10 kg were suitable to transport the full vaccine delivery containing 500 vials for one way delivery.

Table 1 presents the comprehensive calculations of flight times required to transport vaccine to Baduy village through air route with a distance of 1.5 km. The required flight times were different following the variation of drone speeds. Considering the distance to access Baduy village that is constant, each flight delivery time can be calculated according to numbers of transported vials. All delivery times required for vaccine transportation were still below the max flight time of each assessed drone. For full delivery with 500 vials, it requires in average of 3.13 minute delivery time (Figure 5).

**Table 1.** Drone specifications

| Types | Load (kg) | Flight time (max) | Capacity (vials) | Speed (km/h) | Flight time required (min) |
| --- | --- | --- | --- | --- | --- |
| Type 1 | 8 | 20 | 250 | 54 | 1.66 |
| Type 2 | 8 | 18 | 250 | 57 | 1.57 |
| Type 3 | 7 | 15 | 250 | 57 | 1.57 |
| Type 4 | 6 | 16 | 100 | 65 | 1.38 |
| Type 5 | 3 | 50 | 100 | 80 | 1.12 |
| Type 6 | 20 | 16 | 500 | 50 | 1.8 |
| Type 7 | 10 | 16 | 500 | 56 | 1.6 |
| Type 8 | 12 | 30 | 500 | 15 | 6 |

**Figure 5.**
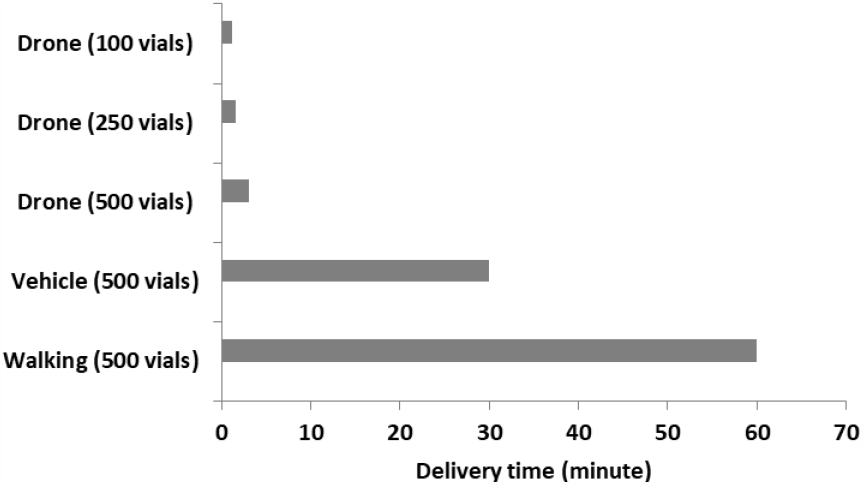
Vaccine required delivery times among various mode transportations and vials.

## 4. Discussions

Logistics and deliveries especially fragile cargo like COVID 19 vaccines trough SE Asia landscaped were very difficult and this can threatened the cargo. Mountainous landscapes where most indigenous people living are frequently experienced landslides (Sidle et al. 2006). Various hillslope positions due to topography (surface and subsurface) and continuity of soil hydrologic pathways (Anderson and Kneale 1982; Fernandes et al. 1994; Tsuboyama et al. 2000) have increased the landslide frequencies in mountainous landscapes. These threats were becoming more imminent mainly during rainy seasons with high rainfall. This weather induced threat combined with the SE Asia rural landscape have illustrated that land network for supporting vaccine delivery is very difficult or even almost impossible.

Studies on COVID 19 logistic have been discussed by Castonguay et al (2020) and Leithäuser et al. (2020). In their study Leithäuser et al. (2020) have emphasized that travel distance and travel restrictions (Castonguay et al. 2020) are several important factors needs to be considered during vaccination. The airline travel distance for one-way travel journey by around 35 km to minimize the barriers for people to get vaccinated. In this study, the distance is not the major issues since the distance is fixed.

Whereas considering the landscape and rugged terrain, the travel time required for delivering the vaccine and implementing the vaccination is the major issue. Travelling by walking requires at least 1 hour for delivering 10 kg payload containing vials. The vaccine cooler box needs to be carried by at least 2 persons. In some areas, there are threats of landslides and even flash flood since the trails are passing wide rivers. Travelling by vehicle only requires 30 minutes whereas this is not the option since the trail width is only 1.5 m and no vehicle can pass it. Delivery of medical goods in times of critical need is not suitable if only supported by wheeled motor vehicles that can be costly and slow (Amukele et al. 2015, Güner et al. 2017; Fancher et al. 2017, Scott & Scott 2018, Thiels et al. 2015).

In this study, the drone delivery was only tested to transport to single point of delivery. Whereas it is possible that the drone will cater the multi point delivery. Berman (2017) has confirmed that a single drone was very effective and can cater 19 delivery points with a 30 minute flight range carrying 2 - 4 kg payload.

Problems that may be encountered during drone delivery is related to the terrain of landing point. Drone assisted vaccine delivery in Vanuatu has reported limited available no flat surface area in the landing area (Martin & Kadura 2019). While this is not an issue in Baduy village since Baduy village was located in valley and field for landing area was available for drone and vaccine landings.

One important factor during vaccine delivery is the environmental factor includes high temperatures and exposure to vibration/agitation. Alteration of vaccine structure due to instability of those factors will impair vaccine biological efficacy. Delivering the vaccine by walking passing though rugged terrain has more exposure on vibration and this will risk the vaccine quality. A comprehensive study on impact of drone assisted delivery on vibration and related vaccine quality has been provided by Hii et al. (2019). Based on their study, a 460 g drone generated 0.1 Hz when landing and high vibration equals to 3.4 Hz was observed when flown forward. When this 3 Hz vibration frequency was tested to the vaccine for 30 minutes, the vibration shows no effect on vaccine quality.

If the vibration is the function of the size of the drone then use of heavy lift drone in this study might be experienced vibration and affect the quality of drones. To anticipate the vibration effects on vaccine quality, there are several approaches that can be taken. The vibrations generated by UAV quadcopter could be minimized by increasing the number of rotors in order to increase stability and the performance of the blades must be tuned accurately. The next option is by using the gimbal technology commonly used for camera mounts.

Regarding the cost required for drone assisted vaccine delivery, Haidari et al. (2016) have provided a comprehensive calculations It is confirmed that implementing the drone in the baseline scenario can improve vaccine availability and even can reduce logistics cost equals to $0.08 per dose vaccine administered with ranges of $0.05-$0.21 per vaccine dose administered.

## 5. Conclusions

Currently, there is an increasing interest in using of drone to deliver medicines to areas that outreach by walking or vehicle delivery (Karaca et al. 2018). Whereas, limited study has been reported about the feasibility and impact of drone in particular massive COVID 19 vaccine delivery that require heavy lift drones. To the best of the authors’ knowledge, this is the first empirical evidences of drone feasibility in supporting COVID 19 vaccine delivery and this study has shed the light that drone transportation of COVID 19 vaccine is feasible from the point of view of effectiveness and efficiency. Then, this study serves as a model for future studies to expand and increase COVID 19 vaccine coverage and delivery using different drones.

## 6. Recommendations

The limitations of this study are only tested vaccine delivery for covering short distance below 10 km though the terrain itself is quite difficult to be accessed by conservative transportation modes. As stated, this study was fit for the purpose of evaluating vaccine transport between rural health facilities to the nearest but isolated tribal villages that village’s access was impeded by forests, hills and rivers. Though to improve and anticipate future COVID 19 vaccine demands by more isolated tribes living deep in the forest within the distance >10 km from the nearest health facilities, future work is recommended to assess the potential use of fixed-wing UAS with capability of vertical take off landing (VTOL)that operate over much larger distances.

## Data Availability

Data available in methodology

